# Higher Comorbidities and Early Death is Characteristic of Hospitalized African-American Patients with COVID-19

**DOI:** 10.1101/2020.07.15.20154906

**Authors:** Raavi Gupta, Raag Agrawal, Zaheer Bukhari, Absia Jabbar, Donghai Wang, John Diks, Mohamed Alshal, Dokpe Yvonne Emechebe, F. Charles Brunicardi, Jason M. Lazar, Robert Chamberlain, Aaliya Burza, M.A. Haseeb

## Abstract

**Background:** African-Americans/Blacks have suffered higher morbidity and mortality from COVID-19 than all other racial groups. This study aims to identify the causes of this health disparity, determine prognostic indicators, and assess efficacy of treatment interventions.

**Method:** We performed a retrospective cohort study of clinical features and laboratory data of COVID-19 patients admitted over a five-week period at the height of the pandemic in the United States. This study was performed at an urban academic medical center in New York City, declared a COVID-only facility, serving a majority Black population

**Result:** Of the 1,070 consecutive patients who tested positive for COVID-19, 496 critically ill patients were hospitalized and included in the study. 88% of patients were Black; and a majority (53%) were 61-80 years old with a mean body mass index in the “obese” range. 97% had one or more comorbidities. Hypertension was the most common (84%) pre-existing condition followed by diabetes mellitus (57%) and chronic kidney disease (24%). Patients with chronic kidney disease and end-stage renal disease who received hemodialysis were found to have significantly lower mortality, then those who did not receive it, suggesting benefit from hemodialysis (11%, OR, 0.35, CI, 0.17 - 0.69 P=0.001). Age >60 years and coronary artery disease were independent predictors of mortality in multivariate analysis. Cox Proportional Hazards modeling for time to death demonstrated a significantly high ratio for COPD/Asthma, and favorable effects on outcomes for pre-admission ACE inhibitors and ARBs. CRP (180, 283 mg/L), LDH (551, 638 U/L), glucose (182, 163 mg/dL), procalcitonin (1.03, 1.68 ng/mL), and neutrophil / lymphocyte ratio (8.5, 10.0) were predictive of mortality on admission and at 48-96 hrs. Of the 496 inpatients, 48% died, one third of patients died within the first three days of admission. 54/488 patients received invasive mechanical ventilation, of which 87% died and of the remaining patients, 32% died.

**CONCLUSIONS:** COVID-19 patients in our predominantly Black neighborhood had higher mortality, likely due to higher prevalence of comorbidities. Early dialysis and pre-admission intake of ACE inhibitors/ARBs improved patient outcomes. Early escalation of care based on comorbidities and key laboratory indicators is critical for improving outcomes in African-American patients.

## Background

Coronavirus Disease 2019 (COVID-19), caused by infection with Severe Acute Respiratory Syndrome Coronavirus-2, has been declared by the World Health Organization to be a pandemic, with over five million confirmed cases.^1^ New York became the epicenter of the epidemic in the United States, accounting for more than 23% of the total U.S. cases by the end of May, 2020.^2^ Such burden of disease is of particular concern since it disproportionately affects communities with considerable health disparities, where African-Americans and Latinos constitute as much as 53% of the population.^3^

The spectrum of COVID-19 presentation ranges from mild influenza-like illness to life-threatening severe respiratory disease requiring ventilatory support.^3^ Comorbid conditions such as hypertension, diabetes mellitus, pulmonary and heart diseases, and demographic factors have been reported to influence outcomes.^4–6^ However, the relative influence of each of these comorbidities in different patient populations and age strata has not been assessed, leading to variability in management and outcomes. Key decisions in patient management such as the choice of antibiotic, blood pressure goals, and perhaps most importantly, airway management strategies, have remained variable across or within hospitals.

National health statistics have documented extensive health disparities for Black COVID-19 patients. They suffer a three-fold greater infection rate, and a six-fold greater mortality rate than their white counterparts.^7^ However, clinical features and laboratory data of prognostic significance from Black COVID-19 patients remain unexplored and undocumented. A range of cultural, linguistic, and healthcare access barriers have prevented clinical investigation. Our hospital, located in New York City, serves a predominantly Black population, and being declared a COVID-only facility, we were able to maintain a standard quality-of-care across all COVID-19 patients.

Here we explore the clinical aspects of COVID-19 and its outcomes in Black patients; we also identify comorbidities and demographic factors that help explain the greater mortality from COVID-19 observed in this vulnerable population. This study evaluated clinical signs and symptoms, laboratory indicators, and management strategies to develop a data-driven COVID-19 patient-care approach. We evaluated these factors at admission and at 48-96 hours after admission, as one-third of our patients died within the first few days of admission, and we found early recognition of critical indicators pivotal in assessing risk of mortality. To our knowledge, this is the only study that correlates early laboratory indicators at two critical time points with outcomes. Therefore, our findings are expected to provide an evidence-based resource for physicians to assess patient progress within the first three days of hospitalization to direct patient management decisions.

## Methods

This study analyzed COVID-19 patients hospitalized at the State University of New York (SUNY), Downstate Medical Center, Brooklyn, New York, that was designated a COVID-only facility by the State of New York as of March 4th, 2020. The hospital is located in a majority Black neighborhood with high rates of poverty.^8^ This study was approved by the SUNY Downstate Institutional Review Board [1587476-1].

We examined 496 consecutively admitted COVID-19 patients during a five-week period (March-April) when the hospital was under peak caseload and admission was reserved for critically ill patients. COVID-19 diagnosis was based on clinical presentation and a positive real-time reverse transcriptase polymerase chain reaction (rtPCR) from a nasopharyngeal swab (Xpert Xpress SARS-CoV-2, Cepheid, Sunnyvale, CA). Patients were admitted if deemed to be in respiratory distress (respiratory rate >22 breaths/min and in need of supplemental oxygen to maintain oxygen saturation >92%), were encephalopathic, or were judged sufficiently ill to require hospitalization. Only patients with final clinical outcomes, such as discharge or expiration, were included. Patients with “fluid overload” (due to missed dialysis), pregnancy, or those still hospitalized were excluded from this analysis.

Demographic factors, comorbidities, presenting clinical symptoms, and outcomes (discharge/death) were available for 496 patients. Of these, laboratory data were recorded for 281 patients on admission or within 24 hr of hospitalization, and at a second time point between 48-96 hr post-admission. Pre-admission medications were recorded based on admission medication reconciliation by admitting physicians. Based on self-reported race/ethnicity, patients were grouped into Black and Others (White Hispanic/non-Hispanic and Asian). HIV-positive patients (with low CD4 counts) and transplant recipients were categorized as “immunocompromised”.

Patients were treated with hydroxychloroquine (200 mg twice a day, for five days) and azithromycin (250 mg once a day, for five days). All patients received standard venous thromboembolism prophylaxis with low-molecular weight heparin or direct oral anticoagulants based on their creatinine clearance rate. Patients with elevated D-dimer received a full dose anticoagulation regimen. Hypoxia, a sign of Acute Respiratory Distress Syndrome (ARDS), was monitored by a continuous pulse oximeter and with arterial blood gas measurements, and supplemental oxygen was provided as needed via noninvasive ventilation. Patients with worsening respiratory distress despite supportive care, as determined by declining pulse oximeter saturation, increasing respiratory rate, or worsening partial pressure of arterial oxygen/percentage of inspired oxygen ratio) were intubated and placed on mechanical ventilation. Patients who developed acute kidney injury (AKI), defined as an absolute increase in serum creatinine greater than or equal to 0.3 mg/dL within 48 hr, with oliguria, with or without hyperkalemia and metabolic acidosis, received hemodialysis.

Computational analysis was conducted using R (ver. 3.6.3).^9^ Continuous variables are presented as median and interquartile range (IQR). Categorical variables such as gender or race are presented as number and percent of patients with 95% confidence intervals (CI). Percentages are expressed based on the available data for the subgroup relative to the total available data for that variable.

Parametric variables were evaluated through a Shapiro-Wilk test of normality with a significance cutoff of *P*<0.01. Non-parametric variables were compared using Mann-Whitney rank sum test, with 95% CIs reported. Categorical variables were evaluated using the Fisher exact test, and odds ratios (OR) alongside 95% CIs are presented. All tests were two-tailed and statistical significance was defined as *P*<0.05. No multiple testing correction was applied. A multivariate linear regression analysis was performed on comorbidities and demographic factors for mortality, and ORs with 95% CIs are presented. Cox proportional hazards analysis for time to death was conducted on comorbidities, demographic factors, and pre-admission medications (angiotensin-converting enzyme (ACE) inhibitors and/or angiotensin II receptor blockers (ARBs) and hazard ratios with 95% CIs are presented.

## Results

1,070 patients were tested for COVID-19 over a 5-week period. After excluding 292 patients who tested negative and 282 who were treated as outpatients, 496 inpatients with positive test results and symptoms consistent with COVID-19 were included in this study.

### Demographic Information

The median patient age was 70 years (Interquartile Range (IQR), 60 - 78). A majority of patients were in the age range of 61-80 years (53%, 255/483) and a small minority were <40 years old (5%, 25/483). Mortality rates inversely correlated with patient age, with the highest mortality rate recorded for the >80-year age group (65%, 64/99). 88% of the patients were Black (424/481) and the remaining 12% were Other. No difference in mortality rates were found between the two groups. Male-to-female ratio was 1.17:1, with a higher mortality rate for males (53%, 139/260). The mean BMI of patients was 30 kg/m^2^ (obese) and no correlation with mortality was found. A majority of patients (81%, 157/194) never smoked and, while not statistically significant, mortality rate increased with any history of smoking (Table 1).

**Table 1.** Demographic Characteristics of COVID-19 inpatients. Demographic characteristics of patients admitted for treatment. The *P value* is calculated between patients who survived and did not survive. BMI, body-mass index; CI, confidence interval

| Variable | Patients | Survivors | Non-survivors | Odds Ratio (95% CI) | P value |
| --- | --- | --- | --- | --- | --- |
| Age - median | 70 | 66 | 73.5 | NA (-10.00 - -5.00) | <0.001 |
| Age ranges | No./total no. (%) | no. (%) | no. (%) |  |  |
| +80 yr. | 99/483(20) | 35 (35) | 64 (65) | 2.20 (1.36 - 3.60) | <0.001 |
| 71-80 yr. | 131/483(27) | 53 (40) | 78 (60) | 1.76 (1.15 - 2.71) | <0.001 |
| 61-70 yr. | 124/483(26) | 63 (51) | 61 (49) | 0.99 (0.64 - 1.52) | 0.84 |
| 51-60 yr. | 66/483(14) | 46 (70) | 20 (30) | 0.39 (0.21 - 0.71) | <0.001 |
| 41-50 yr. | 38/483 (8) | 27 (71) | 11 (29) | 0.39 (0.17- 0.84) | 0.003 |
| 0-40 yr. | 25/483 (5) | 21 (84) | 4 (16) | 0.18 (0.04 - 0.55) | <0.001 |
| Race/Ethnicity | no./total no. (%) | no. (%) | no. (%) |  |  |
| Black | 424/481 (88) | 218 (51) | 206 (49) | 0.73 (0.40 - 1.33) | 0.32 |
| Others | 57/481 (12) | 25 (44) | 32 (56) | 1.35 (0.74 - 2.47) | 0.32 |
| Sex | no./total no. (%) | no. (%) | no. (%) |  |  |
| Male | 260/482 (54) | 121 (47) | 139 (53) | 1.45 (0.99 - 2.11) | 0.44 |
| Female | 222/482 (46) | 124 (56) | 98 (44) | 0.68 (0.47 - 1.00) | 0.44 |
| BMI mean | 30 | 31 | 29 | NA (-2.60 - 1.00) | 0.40 |
| BMI | no./total no. (%) | no. (%) | no. (%) |  |  |
| <29.9 | 132/235 (56) | 45 (34) | 87 (66) | 1.32 (0.75 - 2.34) | 0.34 |
| >30 | 103/235 (44) | 42 (40) | 61 (60) | 0.75 (0.43 - 1.32) | 0.34 |
| Smoking Status | no./total no. (%) | no. (%) | no. (%) |  |  |
| Non-smoker | 157/193 (81) | 80 (51) | 77 (49) | 0.73 (0.33 - 1.60) | 0.46 |
| Past/current smoker | 36/193(19) | 15 (42) | 21 (58) | 1.46 (0.66 - 3.30) | 0.35 |

### Presenting Signs and Symptoms, Comorbidities, and Pre-admission Medication

Presenting patient complaints grouped based on systemic symptoms were “fever” (42%, headache, weakness, lethargy), respiratory (76%, cough, shortness of breath), gastrointestinal (21%, diarrhea, vomiting), and neurological (16%, altered mental status, seizure, unresponsiveness).

Comorbidities were present in 97% (484/496) of patients. The most common comorbidities were hypertension (HT) (84%, 406/484) and diabetes mellitus (DM) (57%, 276/484), followed by chronic kidney disease (CKD)/end stage renal disease (ESRD) (24%,117/485), hyperlipidemia (16%, 81/496), history of cancer (9%, 44/482), coronary artery disease (CAD) (8%, 40/496), chronic obstructive pulmonary disease (COPD) (7%, 30/451), and asthma (6%, 28/449). These comorbidities showed correlation with increased mortality except for HT. Autoimmune diseases (38/482) did not affect outcomes (Table 2). Patients with CKD/ESRD and on dialysis (11%, 52/485) showed lower mortality (*P*=0.001) than counterparts with CKD/ESRD without dialysis (13%, 65/485). These results are notable considering patients with CKD/ESRD suffered higher mean number of comorbidities (mean 4.2) than other patients (mean 3.3, *P* = <0.001) (Table 3).

**Table 2.** Comorbidities in COVID-19 inpatients. Comorbidities among patients admitted for treatment. The *P value* is calculated between patients who survived and did not survive. CKD, chronic kidney disease; COPD, chronic obstructive pulmonary disease; ESRD, end-stage renal disease

| **Comorbidities** | **All Patients**<br>**no./total (%)** | **Survivors**<br>**no. (%)** | **Non-**<br>**survivors**<br>**no. (%)** | **Odds Ratio**<br>**(95% CI)** | **P value** |
| --- | --- | --- | --- | --- | --- |
| **Asthma** | 28/449 (6) | 9 (32) | 19 (68) | 2.44 (1.02 - 6.27) | 0.03 |
| **Autoimmune disease** | 38/482 (8) | 22 (58) | 16 (42) | 0.77 (0.36 - 1.59) | 0.50 |
| **History of cancer** | 44/482 (9) | 14 (32) | 30 (68) | 2.52 (1.25 - 5.29) | <0.001 |
| **COPD** | 30/451 (7) | 14 (47) | 16 (53) | 1.32 (0.58 - 3.01) | 0.057 |
| **Coronary Artery Disease** | 40/496 (8) | 10 (25) | 30 (75) | 3.47 (1.60 - 8.16) | <0.001 |
| **Congestive Heart Failure** | 22/496 (4) | 13 (59) | 9 (41) | 0.72 (0.26 - 1.86) | 0.51 |
| **CKD without dialysis** | 65/485 (13) | 24 (37) | 41 (63) | 2.01 (1.14 - 3.61) | 0.011 |
| **CKD/ESRD with dialysis** | 52/485 (11) | 38 (73) | 14 (27) | 0.35 (0.17 - 0.69) | 0.001 |
| **Diabetes mellitus** | 276/484 (57) | 132 (48) | 144 (52) | 1.48 (1.01 - 2.17) | 0.03 |
| **Hyperlipidemia** | 81/496 (16) | 34 (42) | 47 (58) | 1.57 (0.94 - 2.63) | 0.06 |
| **Hypertension** | 406/484 (84) | 207 (51) | 199 (49) | 1.31 (0.78 - 2.21) | 0.32 |
| **Immune suppression** | 40/475 (8) | 26 (65) | 14 (35) | 0.55 (0.25 - 1.13) | 0.09 |
| **All patients**<br>**≥ 1 Comorbidities** | 484/496 (97) | 252/255 (99) | 232/241 (96) | - | - |

**Table 3.**
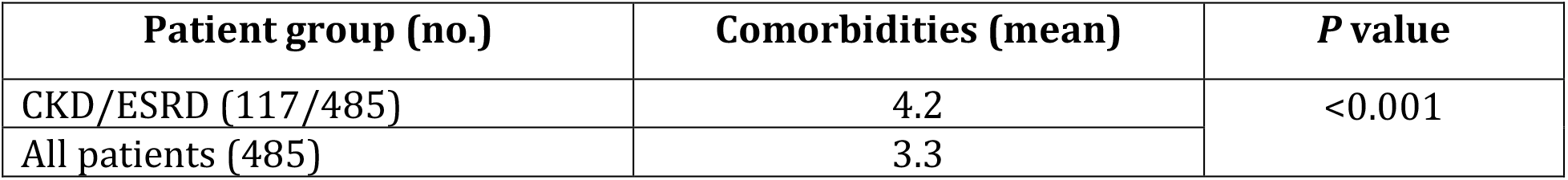
Mean number of comorbidities in patients with chronic kidney diseases and/or end-stage renal disease relative to all patients. Patients with CKD and/or ESRD had a higher mean number of comorbidities than all patients. CKD, Chronic kidney disease; ESRD, end-stage renal disease.

| Patient group (no.) | Comorbidities (mean) | P value |
| --- | --- | --- |
| CKD/ESRD (117/485) | 4.2 | <0.001 |
| All patients (485) | 3.3 |  |

In multivariate analysis, age >60 years and CAD were independent predictors of mortality. Hemodialysis in patients with CKD/ESRD was an independent predictor of lower mortality (*P =* 0.007) (Figure 1). Cox proportional hazards analysis for time to death showed that COPD/Asthma (HR: 1.79, *P* =0.005) had a significantly higher hazards ratio for death (Figure 2). Additional analysis demonstrated patients on pre-admission ACE inhibitors (20%, 29/142) and ARBs (25%, 35/142) had a beneficial effect (*P* = 0.013 *and* 0.036 respectively).

**Figure 1.**
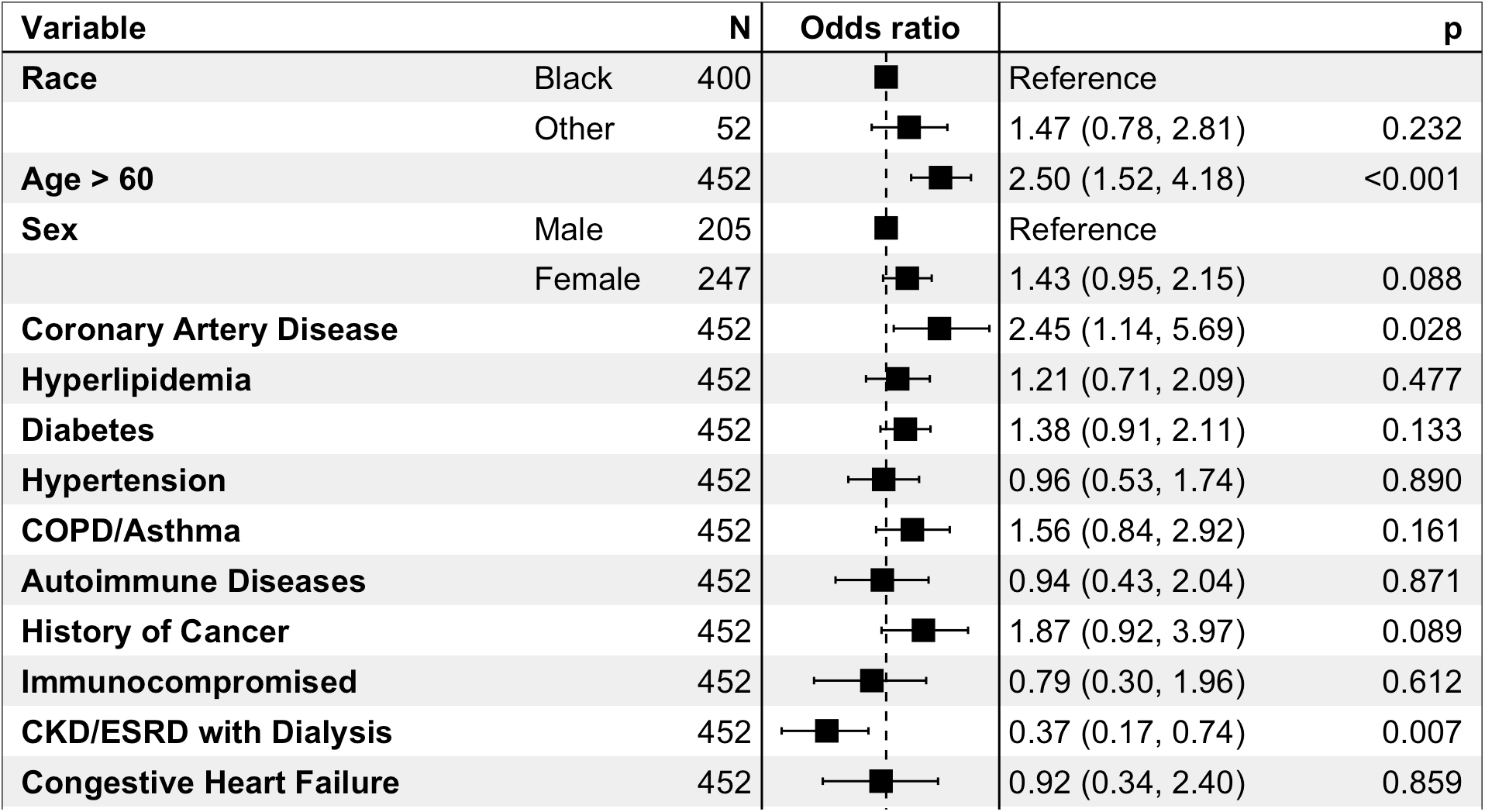
Multivariate logistic regression analysis of the demographic characteristics and comorbidities for mortality. The 95% confidence intervals (CIs) of the odds ratios have not been adjusted for multiple testing and thus do not suggest definitive effects. BMI, body-mass index; CKD, chronic kidney disease; COPD, chronic obstructive pulmonary disease; ESRD, end-stage renal disease.

**Figure 2.**
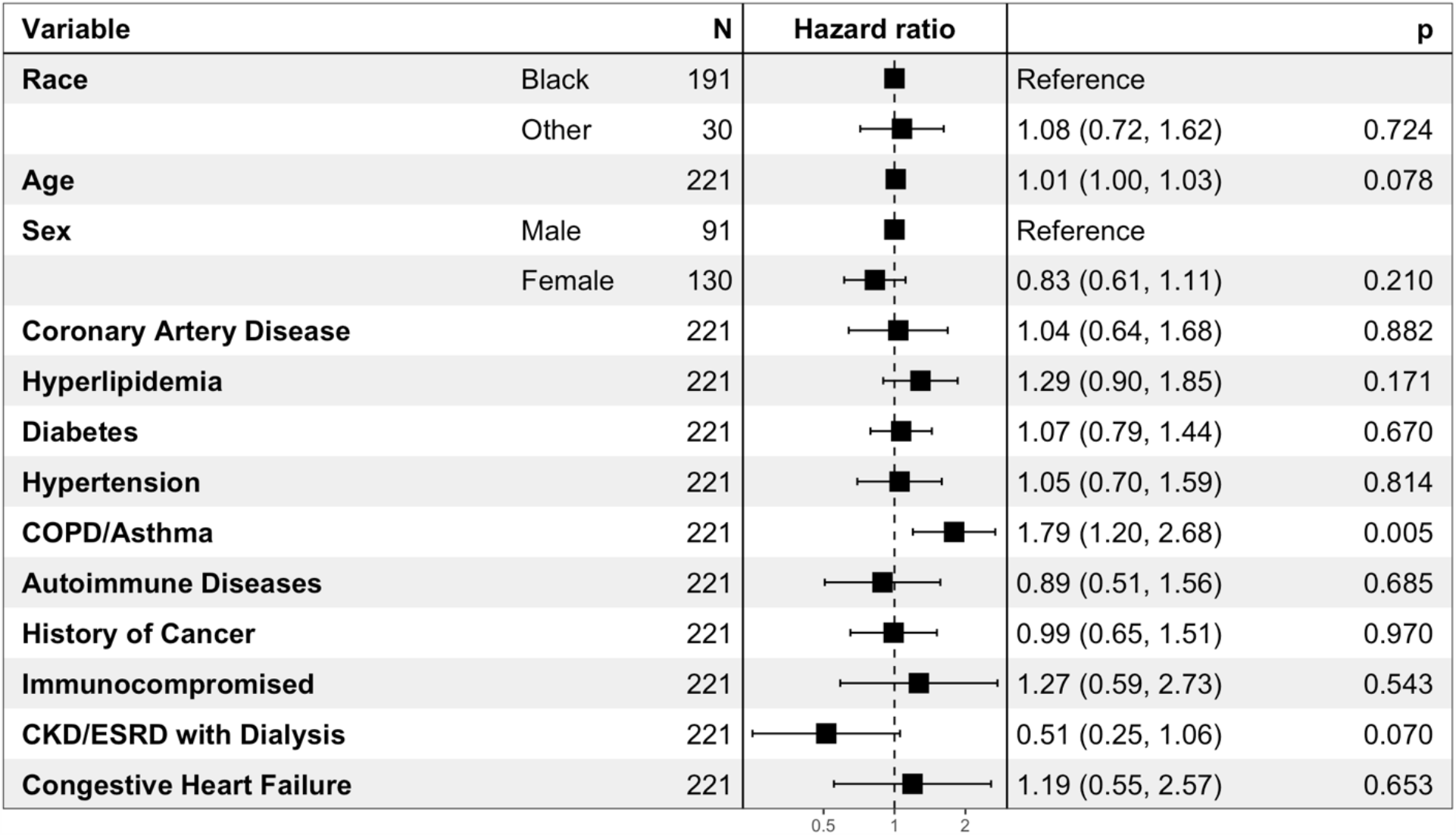
Cox Proportional Hazards analysis on time to death, performed on demographic characteristics and comorbidities.

Complications during clinical course in 312 patients were: acute hypoxic respiratory failure (37%), AKI (15%), cardiogenic shock (18%), neurological shock (5%), sepsis (4%), and diabetic ketoacidosis (3%).

### Laboratory Data

At admission and at 48-96 hr, leukocyte (8.7K/μL, 10.6K/μL) and neutrophil counts (7.3K/μL, 8.9K/μL) were higher (*P*<0.001) and lymphocyte counts (0.8K/μL) were lower at 48-96 hr (*P=*0.003) for non-survivors. The median neutrophil:lymphocyte ratio (NLR) was higher both at admission and at the second time point in patients who did not survive (8.5,10, *P*<0.001). Platelet and hemoglobin were marginally decreased but were not significantly different in survivors and non-survivors. Blood urea nitrogen (BUN) (33, 39 mg/dL), creatinine (1.7, 1.6 mg/dL), glucose (182, 163 mg/dL), alkaline phosphatase (66, 75 U/L), and aspartate aminotransferase (AST) (52, 64 μ/L) levels were higher in non-survivors at both time points (*P*<0.001). Bilirubin and total protein were mildly increased in non-survivors, but were within their respective reference ranges. Albumin (3.4, 2.8 g/dL) was lower for non-survivors at both time points (P<0.001). Lactate dehydrogenase (551, 638 U/L), C-reactive protein (180, 283 mg/L), and procalcitonin (1.03, 1.68 ng/mL) showed significantly higher serum levels at admission and at 48-96 hr (*P* = <0.05) for non-survivors. D-dimer (3.0 mcg/mL, 7.5 times elevation), prothrombin time (PT) (17.2 sec), and international normalized ratio (1.4 U) were increased in non-survivors at the second time point (*P*<0.05). Activated partial thromboplastin time (aPTT) was not found to be different in the two groups (Table 4).

**Table 4.**
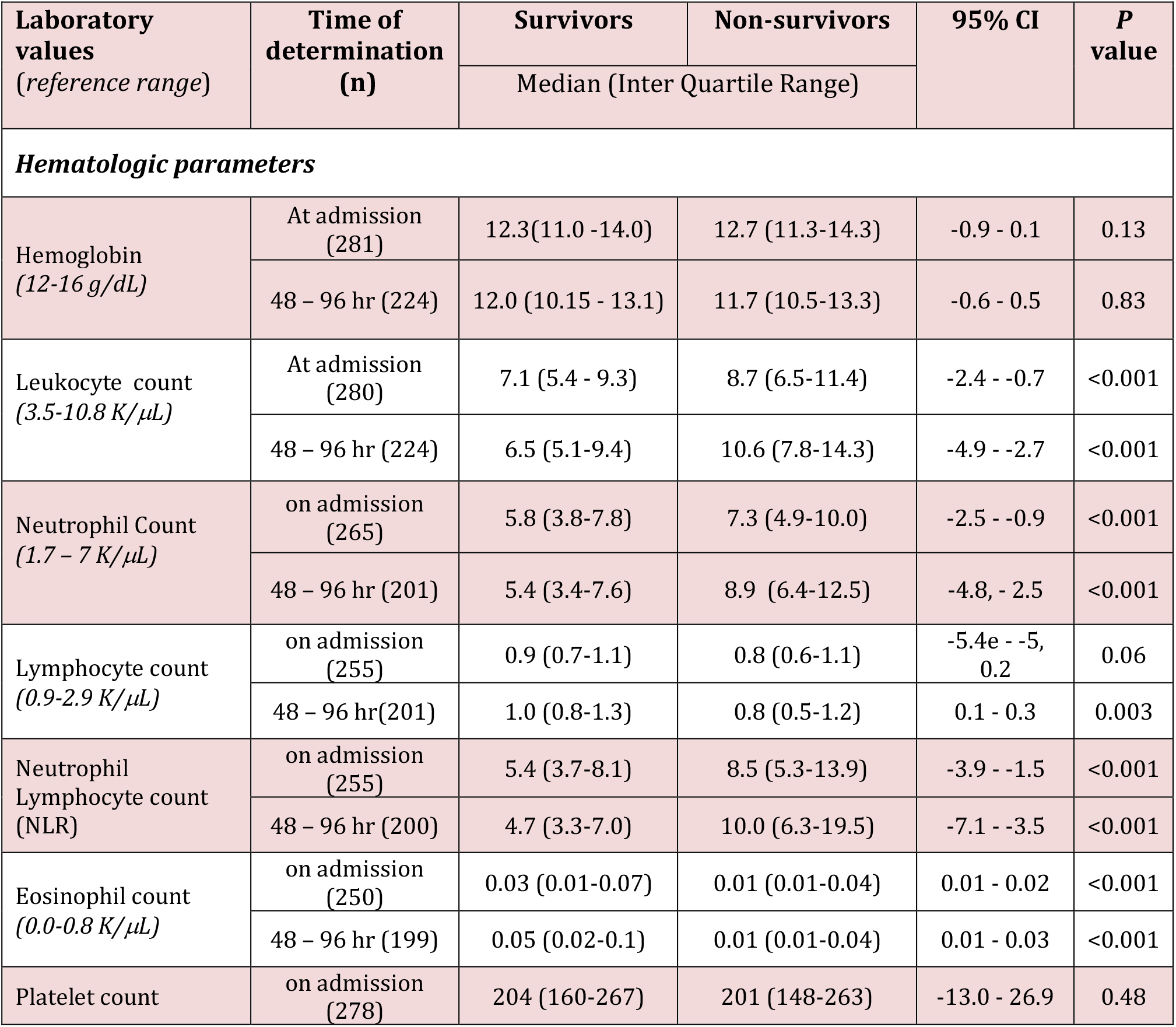
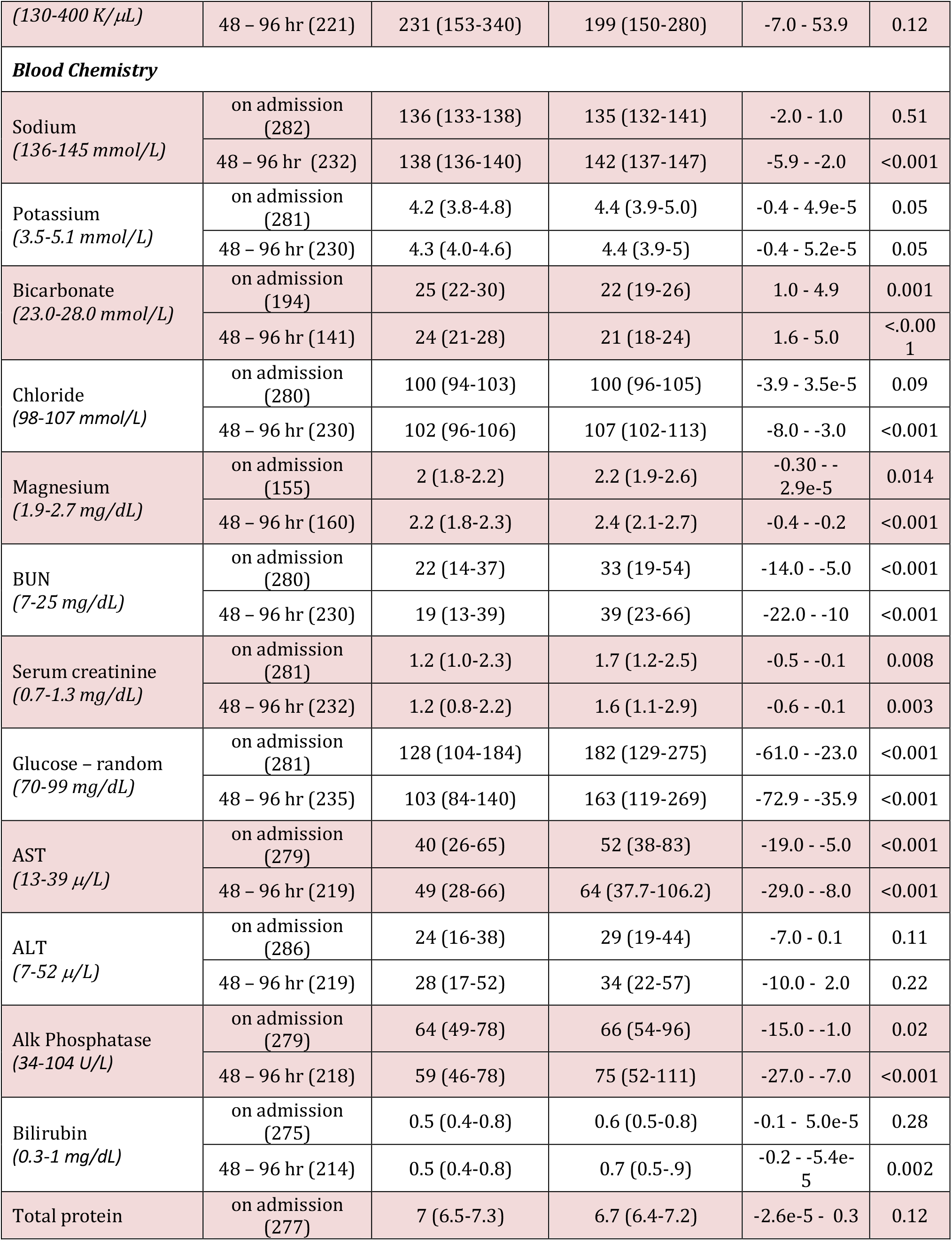
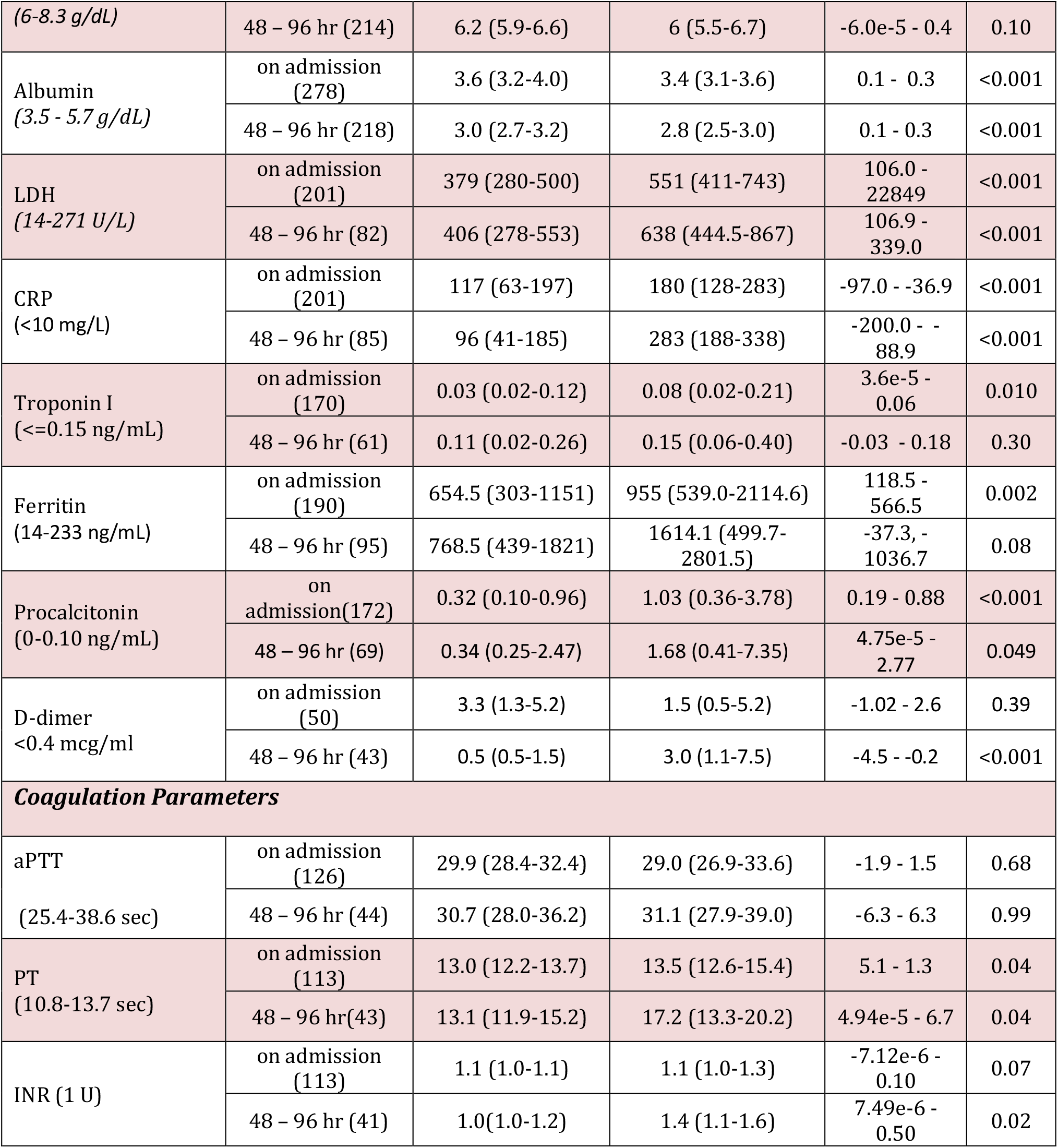
Laboratory Data at admission and at a secondary time point between 48 and 96 hours of admission in hospitalized patients. Laboratory data of 281 inpatients at admission and at a secondary time point between 48 and 96 hours of admission. Median and interquartile ranges are presented. The *P value* is calculated between patients who survived and did not survive. aPTT, activated partial thromboplastin time; Alk Phosphatase, alkaline phosphatase; ALT, alanine aminotransferase; AST, aspartate aminotransferase; BUN, blood urea nitrogen; CI, confidence interval; CRP, C-reactive protein; INR, international normalized ratio; LDH, lactate dehydrogenase; PT, prothrombin time.

### Outcomes

Of the 496 hospitalized patients evaluated, 255 survived and 241 (48.5%) died by the end of the study. Of 488 patients examined, 154 received invasive mechanical ventilation, of which 134 (87%) died. Patients who received supplemental oxygen therapy via non-invasive mode suffered a 32% mortality rate (106/334). This also included patients who self-declared “Do Not Intubate” (DNI), “Do not Resuscitate” (DNR) or came to the hospital in severe respiratory distress and died within the first few hours of admission. More than one third of admitted patients died within the first three days of admission (38%, 91/241), which was similar for both Blacks (77/206) and Others (13/32) (Figure 3). Average time to death for ventilated patients was 7.5 days, while for non-ventilated patients it was 6.2 days from admission.

**Figure 3.**
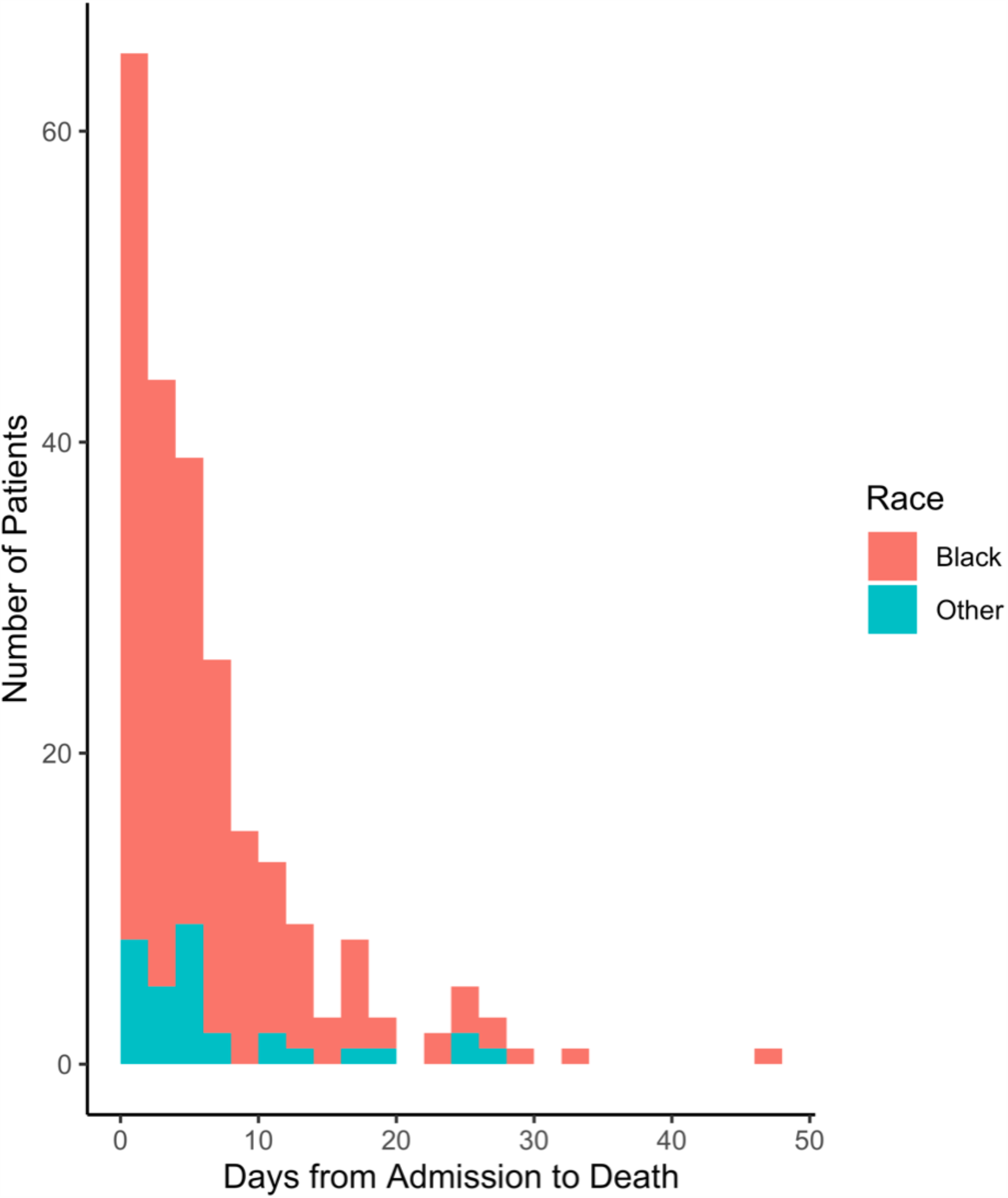
Days from admission to death of 241 consecutive inpatients. More than one third of patients (91/241) died within three days of admission for both Blacks (77/206) and others (13/32). Race was not recorded in 15 patients.

## Discussion

This study documents the demographic, clinical features, and outcomes for patients admitted with COVID-19 at an urban hospital located in an underserved majority-Black neighborhood. We also identify indicators available to physicians at two early time points of evaluation to predict outcomes and develop management plans for appropriate levels of care.

The Black patient population in our study faces unique obstacles such as linguistic and cultural barriers to care and understudied comorbidities.^10,11^ Despite reports that African-Americans face significantly greater mortality from COVID-19, recent studies have examined the clinical outcomes in largely East-Asian or Caucasian cohorts.^10^ Here, we present an analysis of 496 consecutively hospitalized patients who had been diagnosed with COVID-19 over a 5-week period at the height of the pandemic in New York City, and have either been discharged or died.

Older age at admission correlated with higher mortality rate, with the 60+ year age group most at risk, and was an independent risk factor for mortality. Males suffered significantly higher mortality than females, despite identical representation at admission. Recent reports of high plasma concentrations of ACE-2, a receptor for coronavirus, in men may account for higher mortality.^12^ Our inpatient population had a mean BMI in the “obese” range, higher than the national average; this finding mirrors higher BMI amongst the Black population nationwide.^13^ However, BMI was not an independent predictor of survival; higher BMIs were more commonly seen amongst younger patients. Smoking was less prevalent in our patient population than the national average; 4% were current smokers and 15% had quit.^14^ We found smoking to be unrelated to poor outcome.

The majority (88%) of our patients were Black. Race was not an independent prognostic factor for survival; higher mortality in our patient population can be attributed to a greater number and prevalence of comorbidities common amongst this group. Comorbidities were present in 97% of our patients, and the presence of any comorbidity was a strong predictor of mortality, as noted in other recent studies.^15,16^ HT and DM were the two most prevalent preexisting conditions; prevalence of HT (84%) and DM (57%) was considerably higher than previously reported (up to 63% and 36%, respectively).^17–19^ In the multivariate analysis, coronary artery disease was strongly associated with adverse outcome (OR, 2.45, CI, 1.14 - 5.69, *P*=0.034), followed by DM (OR, 1.38, CI, 0.91 - 2.11, *P*=0.13). A 2.5-fold increase in the risk of mortality from COVID-19 in hypertensive patients has been reported, however, this was not discernable in our patients.^18^ Although past history of cancer, HT, autoimmune diseases, and immunosuppression were not independent predictors of mortality, the combined effect of these comorbidities on multiple organ systems and resultant dysregulation of the immune system likely increases susceptibility to COVID-19.^19,20^A notable finding in multivariate analysis was that patients with CKD who were dialyzed early in the course of treatment had better outcomes than those who did not. This group consisted of ESRD patients receiving continued dialysis during hospitalization and patients with worsening CKD who were started early on dialysis (OR, 0.37, CI, 0.17 - 0.74, *P*=0.007). These findings are notable considering patients with CKD/ESRD had more comorbidities as compared to all other patients in the study. Early dialysis stands out as a potentially beneficial treatment option for patients with CKD/ESRD. It is likely that dialysis removes inflammatory mediators, cytokines, and other effector molecules responsible for the end-organ damage. CKD and ESRD were more prevalent in our patient population (24%) than reported in other studies (between 3% to 8.5%), most likely due to complications from HT and DM.^4,21^ Thus, early hemodialysis is critical in Black patients.

We found laboratory data at admission vital for triaging patients to receive intensive care. CRP, LDH, and procalcitonin were significantly increased at both admission and at 48-96 hr in non-survivors. Indicators of AKI, elevated levels of BUN, creatinine, glucose, and reduced levels of bicarbonate or albumin were significant predictors of adverse outcome at both initial and secondary time points. These findings correlate with reported tubular, endothelial, and glomerular capillary loop injury, likely the result of direct injury or systemic hypoxia.^22^ Hypoproteinemia and hypoalbuminemia in non-survivors may result from renal insufficiency and suboptimal nutritional status in critically-ill patients, or could reflect stressed state.^21^ As reported elsewhere, we found hyperglycemia to be a predictor of adverse outcome in COVID-19 patients, regardless of their history of diabetes.^23^ Multivariate analysis of laboratory data was not performed due to sample size limitations.

Peripheral blood analysis showed that a high median NLR at admission and at 48-96 hr was an independent predictor of adverse outcome in COVID-19 patients, as had been reported in other studies.^24^ The presence of COVID-19 associated coagulopathy (CAC), a condition characterized by elevation in fibrinogen and D-dimer levels, high PT, relatively normal aPTT, and mild thrombocytopenia without evidence of microangiopathy, was confirmed in our study.^25^ The mechanisms underlying CAC remain poorly understood, but it can possibly result from activation of extrinsic coagulation pathway, leading to excess consumption of Factor-VII following endothelial cell infection by the virus.^26,27^ Elevated D-dimer levels at the second evaluation time point were associated with higher mortality, likely reflecting coagulation activation from sepsis, “cytokine-storm”, or impending organ failure.

By the end of our 5-week study, 48.5% of the inpatients had died, including 87% of patients who received invasive mechanical ventilation. Reported mortality rates from other retrospective cohort studies ranged from 21% (New York metropolitan area) to 26% (Lombardy region, Italy) and 33% (UK).^4,6,28^ Our mortality rate was elevated relative to other studies, which we believe is due to the largely poor and disadvantaged neighborhood that our hospital serves. Race was not found to be an independent predictor of mortality. The high mortality rate from COVID-19 from this predominantly Black neighborhood demonstrates the large racial disparity in outcomes between these patients and their counterparts elsewhere.

Our study has limitations. It examined a predominantly Black patient cohort, which makes comparisons to other races and ethnicities difficult to quantify. This study was carried out over a five week period at the height of the pandemic in New York City, admissions were restricted to the most seriously ill and hospital resources were under strain, which may have contributed to an increase in overall mortality rates. As knowledge and understanding of COVID-19 was developing during March and early April, complete laboratory studies were not systematically ordered for all patients.

## Conclusions

In our predominantly Black cohort we have recorded a mortality rate from COVID-19 which is significantly greater than that reported in other studies. While race was not an independent predictor of death, this population had a greater burden of comorbidities than the national average and the prevalence of these chronic comorbidities contributed to both disease severity and higher mortality. Our study identified that early escalation of care is important in patients from minority neighborhoods as one third of the admitted patients die within the first three days of admission. Laboratory indicators at admission are predictors of outcome and can be utilized by physicians to triage patients and monitor disease course. As early dialysis in patients with chronic renal insufficiency showed considerable benefit, we believe early dialysis in patients with renal disease can improve outcomes.

## Data Availability

The datasets used and/or analyzed during the current study are available from the corresponding author on reasonable request.

## Abbreviations

ACE: angiotensin converting enzyme
AKI: Acute Kidney Injury
aPTT: activated partial thromboplastin time
Alk Phosphatase: alkaline phosphatase
ALT: alanine aminotransferase
ARB: angiotensin II receptor blockers
ARDS: acute respiratory distress Syndrome
AST: aspartate aminotransferase
BUN: blood urea nitrogen
BMI: body mass index
CAC: COVID-19 associated coagulopathy
CAD: coronary artery disease
CI: confidence interval
CKD: chronic kidney disease
COPD: chronic obstructive pulmonary disease
CRP: C-reactive protein
DM: diabetes mellitus
DNI: do not intubate
DNR: do not resuscitate
ESRD: end stage renal disease
HIV: human immunodeficiency virus
HT: hypertension
INR: international normalized ratio
IQR: interquartile range
LDH: lactate dehydrogenase
NLR: neutrophil:lymphocyte ratio
OR: odds ratio
PT: prothrombin time
rtPCR: real time polymerase chain reaction
SUNY: State University of New York
UK: United Kingdom

## Declarations

### Ethics Approval and Consent to Participate

SUNY Downstate Institutional Review Board [1587476-1].

### Consent for Publication

Not applicable

### Competing Interests

The authors declare that they have no competing interests

### Funding

Not applicable

### Author Contributions

RG and MAH conceived and designed the study. RG, RA, and MAH designed the statistical analysis plan. RG, RA, and MAH analyzed the data and developed the figures and tables. RG, ZB, AJ, DW, JD, MA, and DYE collected data from electronic health records. CFB, JL, RC, and AB provided clinical consultation throughout the study course. All authors contributed intellectual content during the drafting and revision of the work and approved the final version.

## Acknowledgements

Not applicable

## Notes

### Competing Interest Statement

The authors have declared no competing interest.

### Author Declarations

SUNY Downstate Institutional Review Board [1587476-1].

## References

1. WHO Director-General’s opening remarks at the media briefing on COVID-19 - 11 March 2020 [Internet]. [cited 2020 May 23]. Available from: https://www.who.int/dg/speeches/detail/who-director-general-s-opening-remarks-at-the-media-briefing-on-covid-1911-march-2020

2. Cases in the U.S. | CDC [Internet]. [cited 2020 May 16]. Available from: https://www.cdc.gov/coronavirus/2019-ncov/cases-updates/cases-in-us.html

3. U.S. Census Bureau QuickFacts: New York city, New York [Internet]. [cited 2020 May 28]. Available from: https://www.census.gov/quickfacts/newyorkcitynewyork

4. Richardson S, Hirsch JS, Narasimhan M, Crawford JM, McGinn T, Davidson KW, et al. Presenting Characteristics, Comorbidities, and Outcomes Among 5700 Patients Hospitalized With COVID-19 in the New York City Area. JAMA [Internet]. 2020 Apr 22 [cited 2020 Apr 23]; Available from: http://jamanetwork.com/journals/jama/fullarticle/2765184

5. Chen T, Wu D, Chen H, Yan W, Yang D, Chen G, et al. Clinical characteristics of 113 deceased patients with coronavirus disease 2019: retrospective study. BMJ. 2020 26;368:m1091.

6. Docherty AB, Harrison EM, Green CA, Hardwick HE, Pius R, Norman L, et al. Features of 16,749 hospitalised UK patients with COVID-19 using the ISARIC WHO Clinical Characterisation Protocol. medRxiv [Internet]. 2020 Apr 28 [cited 2020 May 23];2020.04.23.20076042. Available from: https://www.medrxiv.org/content/10.1101/2020.0423.20076042v1

7. Yancy CW. COVID-19 and African Americans. JAMA. 2020 Apr 15;

8. East Flatbush Neighborhood Profile [Internet]. [cited 2020 May 23]. Available from: https://furmancenter.org/neighborhoods/view/east-flatbush

9. R: The R Project for Statistical Computing [Internet]. [cited 2020 Apr 23]. Available from: https://www.r-project.org/

10. Dorn A van, Cooney RE, Sabin ML. COVID-19 exacerbating inequalities in the US. The Lancet [Internet]. 2020 Apr 18 [cited 2020 May 15];395(10232):1243–4. Available from: https://www.thelancet.com/journals/lancet/article/PIIS0140-6736(20)30893-X/abstract

11. Laurencin CT, McClinton A. The COVID-19 Pandemic: a Call to Action to Identify and Address Racial and Ethnic Disparities. J Racial Ethn Health Disparities [Internet]. 2020 Apr 18 [cited 2020 May 15];1–5. Available from: https://www.ncbi.nlm.nih.gov/pmc/articles/PMC7166096/

12. Sama IE, Ravera A, Santema BT, van Goor H, ter Maaten JM, Cleland JGF, et al. Circulating plasma concentrations of angiotensin-converting enzyme 2 in men and women with heart failure and effects of renin–angiotensin–aldosterone inhibitors. Eur Heart J [Internet]. 2020 May 14 [cited 2020 May 16];41(19):1810–7. Available from: http://academic.oup.com/eurheartj/article/41/19/1810/5834647

13. Fryar CD, Kruszon-Moran D, Gu Q, Ogden CL. Mean Body Weight, Height, Waist Circumference, and Body Mass Index Among Adults: United States, 1999-2000 Through 2015-2016. Natl Health Stat Rep. 2018;(122):1–16.

14. Creamer MR. Tobacco Product Use and Cessation Indicators Among Adults — United States, 2018. MMWR Morb Mortal Wkly Rep [Internet]. 2019 [cited 2020 May 15];68. Available from: http://www.cdc.gov/mmwr/volumes/68/wr/mm6845a2.htm

15. Wang B, Li R, Lu Z, Huang Y. Does comorbidity increase the risk of patients with COVID-19: evidence from meta-analysis. Aging. 2020 Apr 8;12(7):6049–57.

16. Guan W, Liang W, Zhao Y, Liang H, Chen Z, Li Y, et al. Comorbidity and its impact on 1590 patients with Covid-19 in China: A Nationwide Analysis. Eur Respir J [Internet]. 2020 Jan 1 [cited 2020 May 16]; Available from: https://erj.ersjournals.com/content/early/2020/03/17/13993003.00547-2020

17. Garg S. Hospitalization Rates and Characteristics of Patients Hospitalized with Laboratory-Confirmed Coronavirus Disease 2019 — COVID-NET, 14 States, March 1– 30, 2020. MMWR Morb Mortal Wkly Rep [Internet]. 2020 [cited 2020 May 16];69. Available from: https://www.cdc.gov/mmwr/volumes/69/wr/mm6915e3.htm

18. Lippi G, Wong J, Henry BM. Hypertension in patients with coronavirus disease 2019 (COVID-19): a pooled analysis. Pol Arch Intern Med. 2020 30;130(4):304–9.

19. Leoncini G, Viazzi F, Storace G, Deferrari G, Pontremoli R. Blood pressure variability and multiple organ damage in primary hypertension. J Hum Hypertens. 2013 Nov;27(11):663–70.

20. Restrepo MI, Sibila O, Anzueto A. Pneumonia in Patients with Chronic Obstructive Pulmonary Disease. Tuberc Respir Dis. 2018 Jul;81(3):187–97.

21. Su H, Yang M, Wan C, Yi L-X, Tang F, Zhu H-Y, et al. Renal histopathological analysis of 26 postmortem findings of patients with COVID-19 in China. Kidney Int [Internet]. 2020 Apr 9 [cited 2020 May 16];0(0). Available from: https://www.kidney-international.org/article/S0085-2538(20)30369-0/abstract

22. Huang C, Wang Y, Li X, Ren L, Zhao J, Hu Y, et al. Clinical features of patients infected with 2019 novel coronavirus in Wuhan, China. Lancet Lond Engl. 2020 15;395(10223):497–506.

23. Li X, Xu S, Yu M, Wang K, Tao Y, Zhou Y, et al. Risk factors for severity and mortality in adult COVID-19 inpatients in Wuhan. J Allergy Clin Immunol. 2020 Apr 12;

24. Liu Y, Du X, Chen J, Jin Y, Peng L, Wang HHX, et al. Neutrophil-to-lymphocyte ratio as an independent risk factor for mortality in hospitalized patients with COVID-19. J Infect. 2020 Apr 10;

25. COVID-19 and Coagulopathy - Hematology.org [Internet]. [cited 2020 May 16]. Available from: https://www.hematology.org:443/covid-19/covid-19-and-coagulopathy

26. Ferrario CM, Jessup J, Chappell MC, Averill DB, Brosnihan KB, Tallant EA, et al. Effect of angiotensin-converting enzyme inhibition and angiotensin II receptor blockers on cardiac angiotensin-converting enzyme 2. Circulation. 2005 May 24;111(20):2605–10.

27. Varga Z, Flammer AJ, Steiger P, Haberecker M, Andermatt R, Zinkernagel AS, et al. Endothelial cell infection and endotheliitis in COVID-19. Lancet Lond Engl. 2020 02;395(10234):1417–8.

28. Grasselli G, Zangrillo A, Zanella A, Antonelli M, Cabrini L, Castelli A, et al. Baseline Characteristics and Outcomes of 1591 Patients Infected With SARS-CoV-2 Admitted to ICUs of the Lombardy Region, Italy. JAMA. 2020 06;

